# Secular trends in the incidence, prevalence, and survival of primary liver cancer in the United Kingdom from 2000-2021: a population-based cohort study

**DOI:** 10.1101/2024.08.05.24311466

**Authors:** Berta Cuyàs, Edilmar Alvarado-Tapias, Eng Hooi Tan, Asieh Golozar, Talita Duarte-Salles, Antonella Delmestri, Josep Maria Argemí Ballbé, Wai Yi Man, Edward Burn, Carlos Guarner-Argente, Daniel Prieto Alhambra, Danielle Newby

## Abstract

**Background:** Primary liver cancer (PLC) remains a global health challenge. Understanding trends in the disease burden and survival is crucial to inform decisions regarding screening, prevention and treatment.

**Methods:** Population-based cohort study using UK primary care data from the Clinical Practice Research Datalink (CPRD) GOLD (2000 to 2021), replicated in CPRD Aurum. PLC incidence rates (IR), period prevalence (PP) and survival at one, five and ten years over the study period were calculated, and stratified by age, sex and diagnosis year.

**Results:** The crude IR of PLC was 4.56 (95%CI 4.42-4.70) per 100,000 person-years between 2000 and 2021, with an increase over time across age and sex strata. Sex-specific IR for males was higher than females, 6.60 (95%CI 6.36-6.85) vs. 2.58 (95%CI 2.44-2.74) per 100,000 person-years. Crude PP showed a 7-fold increase over the study period, with PP 0.02% (95%CI 0.019%-0.022%) in 2021, and a 2.8-fold higher PP in males. Survival at one, five and ten years after diagnosis was 41.7%, 13.2% and 7.1%, respectively, for both sexes. One-year survival increased only in men, from 33.2% in 2005-2009 to 49.3% in 2015-2019.

**Conclusion:** Over the past two decades, there has been a significant increase in the number of patients diagnosed with PLC. Despite a slight improvement in median and one-year survival in men, prognosis remains poor. To improve the survival of PLC patients, it is necessary to understand the epidemiological changes and address the preventable risk factors associated with liver disease and promote early detection and access to care.

**LAY SUMMARY:** This population-based cohort study shows that the incidence and prevalence of primary liver cancer in the UK has increased in the last 20 years across both sexes and age groups, with a 7-fold increase in crude period prevalence over the study period. One-year survival has improved only in males over the study period and, regrettably, no increases in long-term survival were observed. Our findings are a call for awareness to stimulate further research and public health actions on liver cancer.

## INTRODUCTION

Primary liver cancer (PLC) is the sixth most common cancer and the third leading cause of cancer death worldwide (1). Hepatocellular carcinoma (HCC) is the most common form of PLC accounting for approximately 90% of cases (2).

The incidence and mortality of PLC are growing worldwide, with approximately 906,000 new cases and 830,000 deaths in 2020. Incidence remains highest in Eastern Asia and Northern Africa, although it is increasing in different parts of Europe, Americas, and Oceania (3). PLC was also one of the top five causes of cancer mortality in some countries in Europe, such as Bosnia and Herzegovina, France, Italy, Republic of Moldova, and Romania; and Western Asia (4). The incidence and mortality from PLC are predicted to rise by more than 50% over the next 20 years (1) with the UK showing a rapid rate of incident cases and projected to have one of the highest annual increases over the next decade (5).

Improvements in survival have been made over previous decades due to advancements in chemotherapy, surgical techniques, and the shift towards multidisciplinary teams to manage care. Despite this, survival is still poor even in high-income countries compared to other cancers with one and five-year relative survival estimates around 40% and 10% respectively (4,6).

The main risk factors linked to PLC are cirrhosis, Hepatitis B virus (HBV), Hepatitis C virus (HCV), harmful alcohol consumption and metabolic factors such as diabetes and obesity (7). With the use of direct-acting antiviral therapy, the risk attributed to HCV has substantially decreased globally, while alcohol-associated liver disease (ALD) and metabolic dysfunction-associated steatotic liver disease (MASLD) are becoming more prominent risk factors for PLC (8). Less prevalent risk factors include autoimmune hepatitis, hemochromatosis, _α_1-antitrypsin deficiency and aflatoxin ingestion. Regarding sociodemographic characteristics, older age, being male and some racial or ethnic minorities (in particular, Hispanics) as well as lifestyle factors such as cigarette smoking have also been associated with HCC (2). The distribution of these risk factors has gradually changed over time and between populations (2).

Understanding trends in the incidence, prevalence, and overall survival of liver cancer is an important aspect to inform decisions regarding screening, prevention, treatment, and disease management. Due to the increases in risk factors for PLC such as obesity, alcohol consumption and diabetes, a comprehensive assessment of the trends and disease burden of PLC is lacking in the UK. We therefore set out to characterise the secular trends of PLC in terms of incidence, prevalence and survival in the UK.

## METHODS

### Data sources and Study design

We carried out a population cohort study using routinely collected primary care data from the UK. People with a diagnosis of PLC and a background cohort (denominator population) were identified from Clinical Practice Research Datalink (CPRD) GOLD (July 2022). We additionally carried out this study using CPRD Aurum to compare the results with CPRD GOLD. These databases contain pseudo-anonymised patient-level information on demographics, lifestyle data, clinical diagnoses, prescriptions, and preventive care contributed by general practitioners from the UK. CPRD GOLD contains data from across the UK whereas Aurum only contains data from England. The use of CPRD data was approved by CPRD’s Research Data Governance process (22_001843). GOLD and Aurum are established primary care databases broadly representative of the UK population (9). Both databases were mapped to the Observational Medical Outcomes Partnership (OMOP) Common Data Model (CDM) (10,11).

### Study participants and time at risk

All individuals were required to be aged 18 years or older and have at least one year of prior history. For the incidence and prevalence analysis, the study cohort consisted of individuals present in the databases from 1st January 2000. For CPRD GOLD, these individuals were followed up to whichever came first: the practice stopped contributing to the database, the patient left the practice, date of death, or the 31st of December 2021 (the end of study period) whereas for Aurum, the end of the study period was 31st of December 2019. For the survival analysis, only individuals with a newly diagnosed cancer were included. These individuals were followed up from the date of their diagnosis to either date of death, practice stopped contributing to the database, patient left the practice, or end of the study period. Any patients whose death and cancer diagnosis occurred on the same date were removed from the survival analysis.

### Primary liver cancer definitions

We used Systematized Nomenclature of Medicine - Clinical Terms (SNOMED CT) diagnostic codes to identify PLC events. Diagnostic codes related to intrahepatic cholangiocarcinoma were excluded. Diagnostic codes indicative of either non-malignant cancer or metastasis were excluded as well as diagnosis codes indicative of melanoma and lymphoma occurring in the organs of interest. The study outcome cancer definition was reviewed with the aid of the Cohort Diagnostics R package (12). This package was used to identify additional codes of interest and to remove those highlighted as irrelevant based on feedback from clinicians with oncology, primary care, and real-world data expertise through an iterative process during the initial stages of analyses. The clinical code lists used to define PLC can be found in supplementary information S1. OMOP-based computable phenotypes are available, together with all analytical code on Github to enable reproducibility (https://github.com/oxford-pharmacoepi/EHDENCancerIncidencePrevalence). For overall and annual crude incidence rates (IR) and annual prevalence, all PLC events in the period 2000-2021 were included. For survival analyses, mortality was defined as all-cause mortality based on date of death records.

### Statistical methods

The population characteristics of patients with a diagnosis of PLC were summarised, with median and interquartile range (IQR) used for continuous variables and counts and percentages used for categorical variables.

For incidence, the number of events, the observed time at risk, and the incidence rate per 100,000 person years were summarised along with 95% confidence intervals (95% CI). Annual crude incidence rates were calculated as the number of incident PLC cases as the numerator and the recorded number of person-years in the general population within that year as the denominator whereas overall incidence was calculated from 2000 to 2021.

Age-standardized IRs were calculated using the 2013 European Standard Population (ESP2013) (21). The ESP2013 is a population standard with a predefined age distribution which accounts for differences in age structures between different populations to ensure fair comparisons. The ESP2013 provides predefined age distribution in five-year age bands; therefore, we collapsed these to obtain distributions for ten-year age bands used in this study. We used the age distribution of 20-29 years from ESP2013 for age-standardization as age distributions were not available for 18-29 years age band used in this study.

Period prevalence was calculated on 1st January for the years 2000 to 2021, with the number of patients fulfilling the case definition for liver cancer as the numerator. The denominator was the participants eligible on 1st January in the respective years for each database. The number of events, and prevalence (%) were summarised along with 95% confidence intervals.

For survival analyses, we used the Kaplan-Meier method to estimate the overall survival probability from observed survival times with 95% confidence intervals. We estimated the median survival and survival probability, one, five, and ten years after diagnosis. Any patients whose death date and cancer diagnosis date occurred on the same date were removed from the survival analysis.

All results were stratified by database, age (ten-year age bands apart from the first and last age bands which were 18-29 years and 90+ years, respectively) and sex. For survival analysis, we additionally stratified by calendar time of cancer diagnosis (2000-2004, 2005-2009, 2010-2014, 2015-2019 and 2020-2021) allowing a maximum of five years follow-up from cancer diagnosis. To avoid re-identification, we do not report results with fewer than five cases.

For Aurum, the same statistical analyses were performed using data from 1st January 2000 to 31st December 2019 to compare with results from GOLD apart from the calendar time stratification which was only performed in GOLD.

The statistical software R version 4.2.3 was used for analyses. For calculating incidence and prevalence, we used the Incidence Prevalence R package (13).

## RESULTS

### Patient Populations and characteristics

Overall, there were 11,388,117 eligible patients, with at least one year of prior history identified from January 2000 to December 2021 from CPRD GOLD. Attrition tables for this study can be found in the supplementary information (Supplement S2). A summary of study patient characteristics of those with a diagnosis of PLC for GOLD is shown in Table 1.

**Table 1:** Baseline characteristics of primary liver cancer patients at the time of diagnosis stratified by sex from CPRD GOLD.

| **Sex** | **Male** | **Female** |
| --- | --- | --- |
| **Number of patients** | 2,848 | 1,151 |
| **Age in years, Median (IQR)** | 70 (62 to 77) | 73 (64 to 80) |
| **Age groups in years, N (%)** |  |  |
| 18-29 | 11 (0.4%) | <5 |
| 30-39 | 18 (0.6%) | 9 (0.8%) |
| 40-49 | 97 (3.4%) | 41 (3.6%) |
| 50-59 | 411 (14.4%) | 149 (12.9%) |
| 60-69 | 811 (28.5%) | 260 (22.6%) |
| 70-79 | 952 (33.4%) | 385 (33.4%) |
| 80-89 | 512 (18.0%) | 271 (23.5%) |
| 90+ | 36 (1.3%) | 33 (2.9%) |
| **Prior history in days, Median (IQR)** | 3,977 (2,189 to 5,642) | 3,943 (2,110 to 5,642) |
| **Comorbid conditions (any time prior)** |  |  |
| **Chronic liver disease, N (%)** | 663 (23.3%) | 176 (15.3%) |
| **Recorded risk factors for PLC, N (%)** |  |  |
| Alcoholic liver disease (ALD) | 385 (13.5%) | 52 (4.5%) |
| Hepatitis C | 91 (3.2%) | 25 (2.2%) |
| Hepatitis B | 20 (0.7%) | <5 |
| Non-alcoholic fatty liver disease (NAFLD) | 80 (2.8%) | 26 (2.3%) |
| Hemochromatosis | 90 (3.2%) | 9 (0.8%) |
| Autoimmune hepatitis | 14 (0.5%) | 37 (3.2%) |
| **Other recorded risk factors, N (%)** |  |  |
| Hypertensive disorder | 850 (29.8%) | 349 (30.3%) |
| Diabetes | 851 (29.9%) | 215 (18.7%) |
| Hyperlipidaemia | 209 (7.3%) | 99 (8.6%) |
| Obesity | 313 (11.0%) | 102 (8.9%) |
| **Smoking Status (any time 5 years prior), N (%)** |  |  |
| Current/former smoker | 806 (28.3%) | 270 (23.0%) |
| Nonsmoker | 954 (33.5%) | 559 (48.6%) |
| Missing/no records | 1,088 (38.2%) | 322 (28.0%) |

There were 3,999 patients with PLC in CPRD GOLD. Overall, those diagnosed with PLC were more likely to be male (71%), with a median age of 71 (IQR 62 to 77) years at presentation. The highest percentage of PLC patients were those aged 70-79 years old contributing to 33.4% of diagnosed patients, for both males and females, with similar observations in Aurum (Supplement S3). Males had higher prevalence of chronic liver disease, ALD and hemochromatosis, diabetes and were more likely to be smokers compared to females. However, females had higher proportions of autoimmune hepatitis compared to males. Similar percentages regarding HCV and NAFLD were observed in PLC patients across both sexes.

### Incidence rates stratified by calendar year, age and sex

The overall crude IR of liver cancer in 2000 to 2021 was 4.56 (95% CI 4.42 to 4.70) per 100,000 person-years. Sex-specific IR for females was 2.58 (95% CI 2.44 to 2.74) and for males was 6.60 (95% CI 6.36 to 6.85) per 100,000 person-years, with similar results in Aurum. Annualised IRs increased from 2000 to 2019 for the whole population and both sexes with males having higher rates (Figure 1). For GOLD, IR dropped in 2020 before bouncing back in 2021. Age standardized incidence rates for CPRD GOLD for both sexes showed similar trends (Supplement S4). All study results for this study can be found and downloaded in a user-friendly interactive web application: https://dpa-pde-oxford.shinyapps.io/LiverCancerIncPrevSurvShiny/.

**Figure 1:**
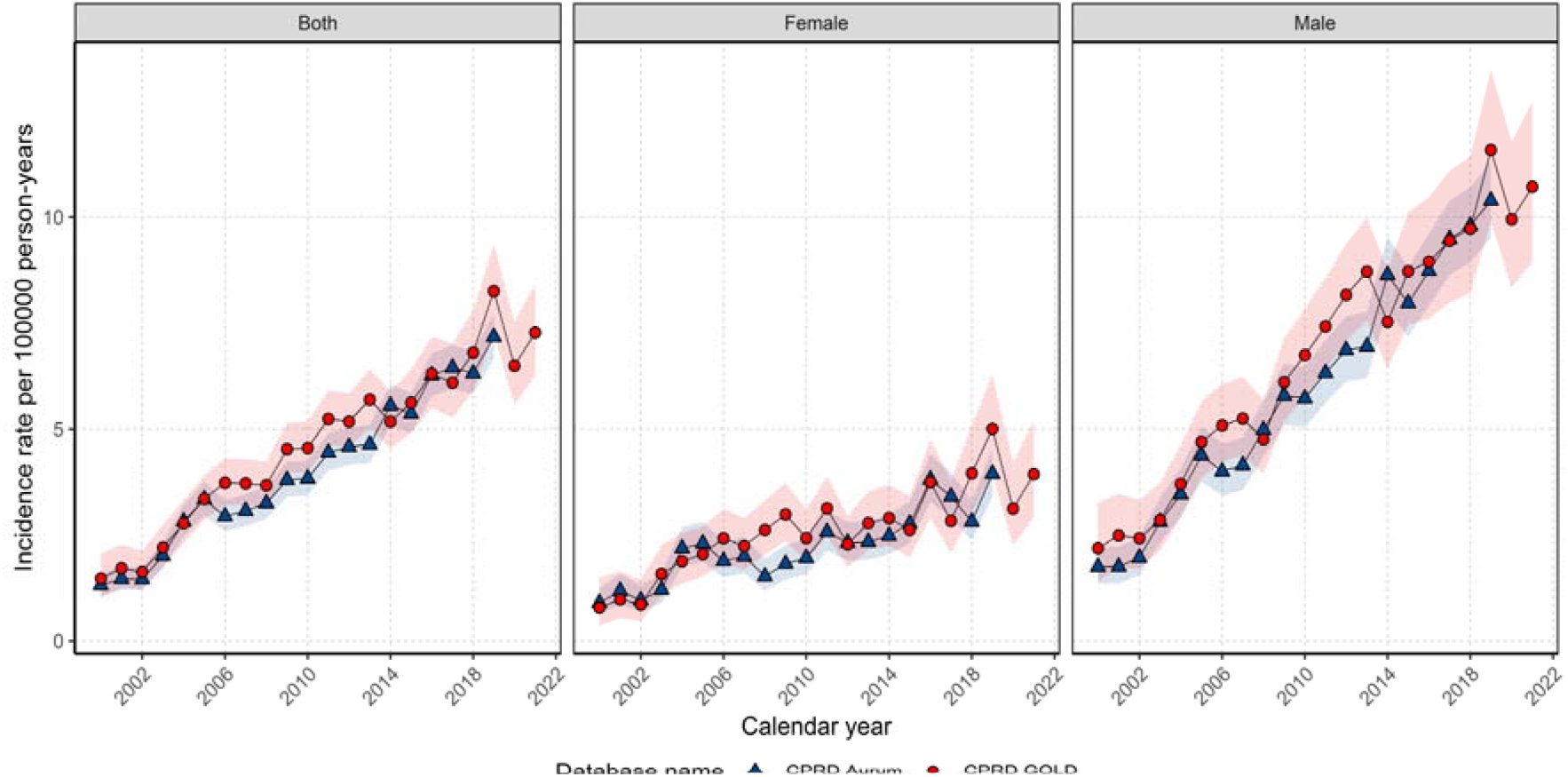
Annual incidence rates for PLC from 2000 to 2021 stratified by database and sex.

Overall crude IRs were higher with increasing age up to 80–89 years. Those aged 18 to 29 had the lowest overall IRs with an IR of 0.09 (95% CI 0.05 to 0.15) per 100,000 person years, whereas those aged 80–89-years had the highest IR of 17.7 (95% CI 16.4-18.9) with similar or slightly lower IRs in Aurum (Supplement S5).

Annualised IRs for each age group (Figure 2) show IRs have increased over the study period for those aged 40-89 years of age. For other age groups there was not enough data to assess annualised IR trends. Stratification on both sex and age showed similar trends to Figure 2 for both sexes (Supplement S6). Males had higher IRs across the study period with the differences in IR between males and females widening over the study period.

**Figure 2:**
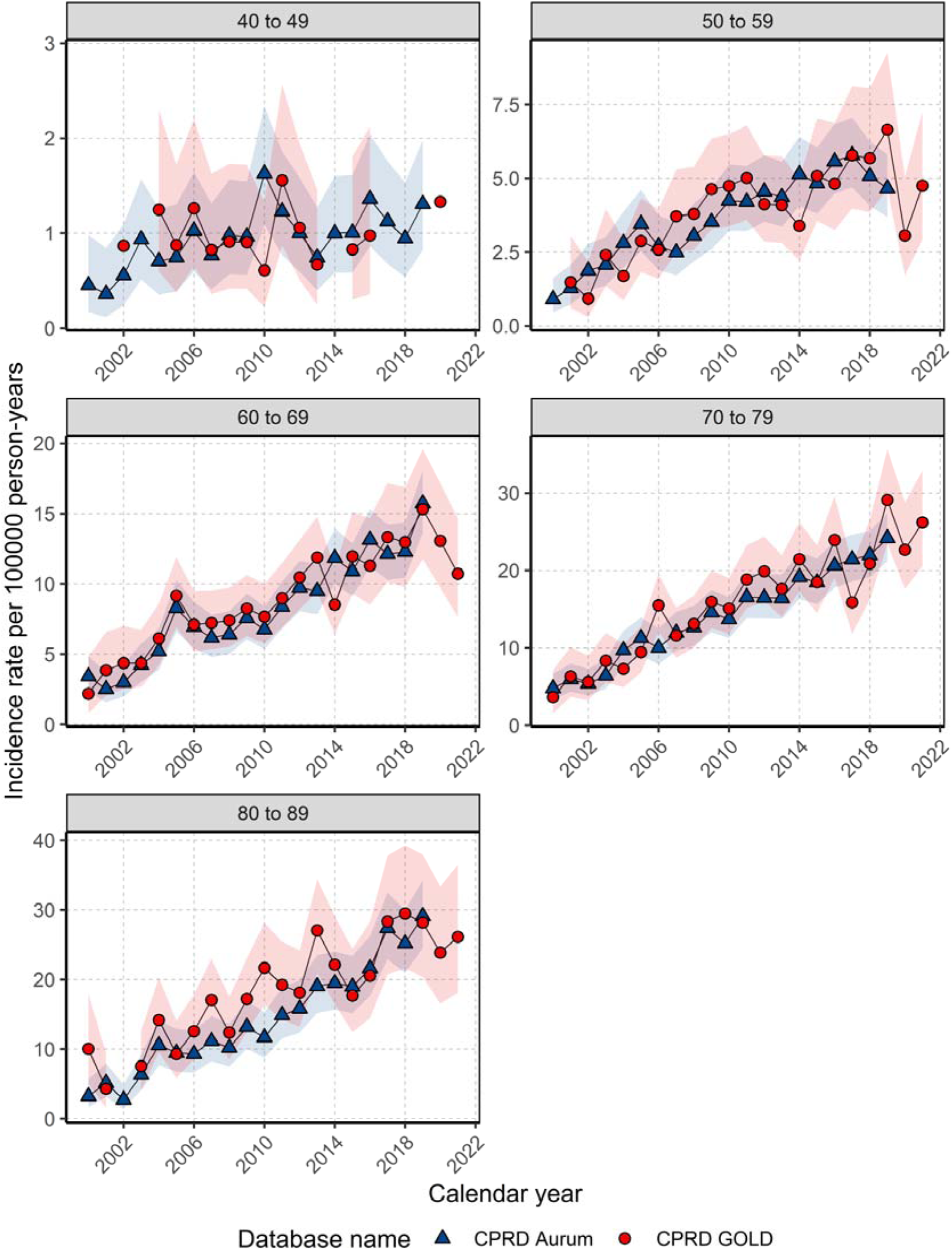
Annualised incidence rates from 2000 to 2021 stratified by database and age group.

### Prevalence for study population with database, age, and sex stratifications

For the whole population in GOLD, crude PP for PLC in 2021 was 0.020% (0.018% to 0.022%) and 2.72-fold higher for males compared to females. Since 2000, PP has increased 6.66-fold across the study period with males seeing a 10-fold increase in PP compared to 5.5-fold increase in females. Similar PP was observed when comparing PP from 2000 to 2019 across both databases (Figure 3).

**Figure 3:**
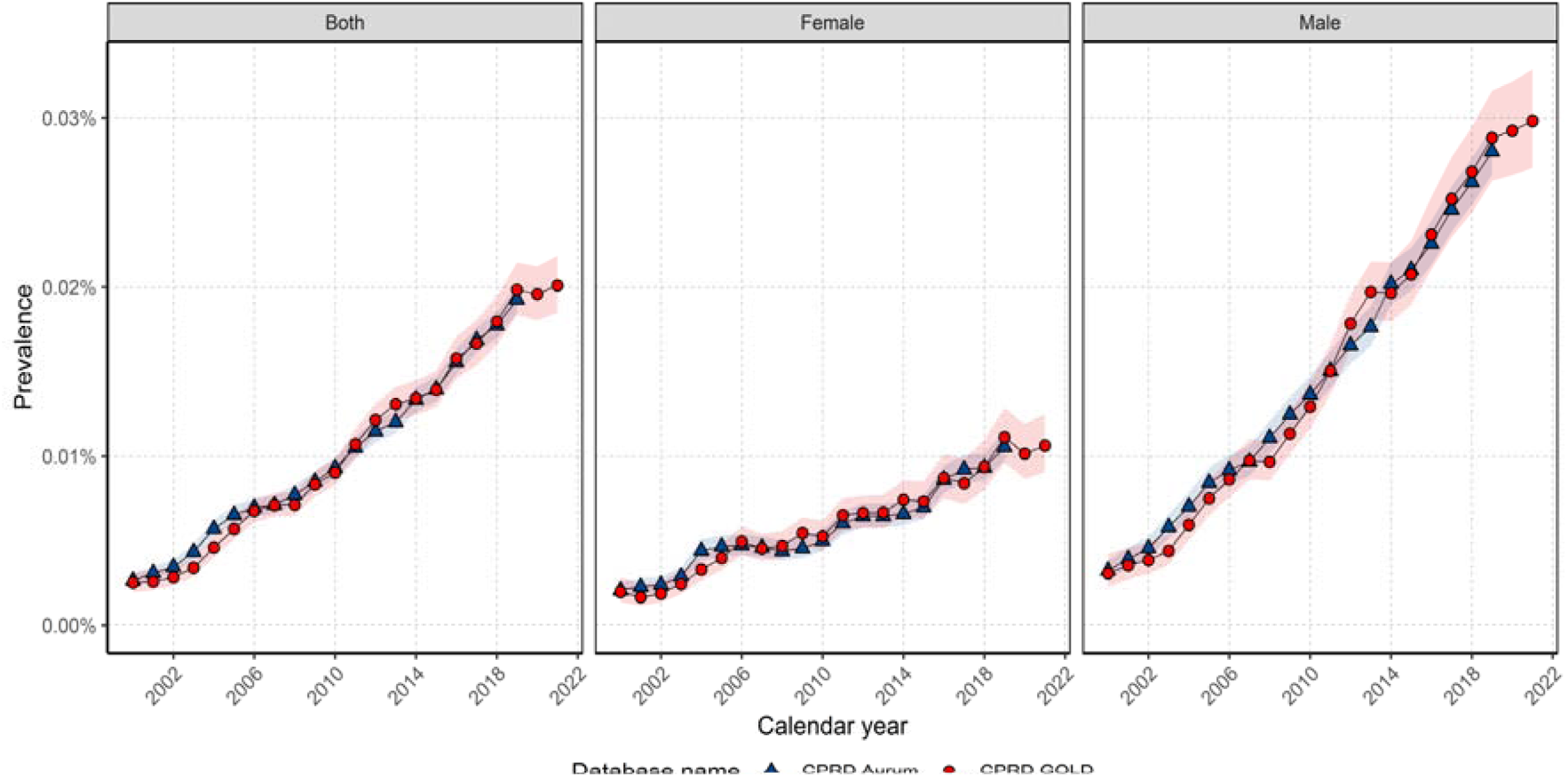
Annual prevalence from 2000 to 2021 for whole population and stratified by sex.

When stratifying by age group, PP in 2021 was highest in those aged between 70-89 years of age (0.07%). PP increased over the study period for all age groups across both databases except for those aged 40-49 in GOLD where there was a little change in PP over time between 2011-2021, whereas Aurum showed a gradual increase in PP over the study period (Figure 4). Stratification on both sex and age group showed similar trends with females driving the increase in PP in those aged 40-49 years old in Aurum (Supplement S7).

**Figure 4:**
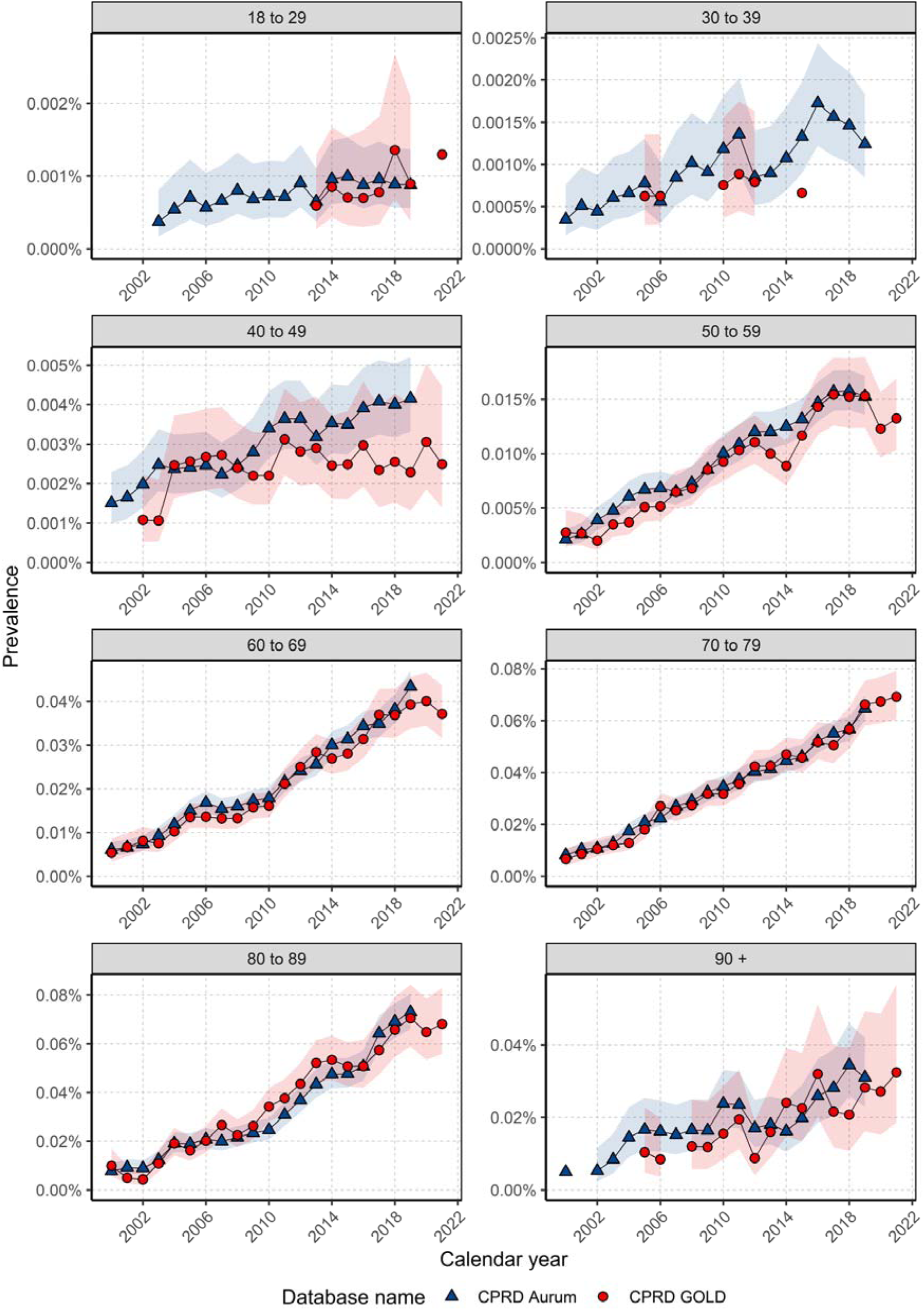
Annual prevalence from 2000 to 2021 stratified by database and age group.

### Overall survival rates for cancer population with age, sex, and calendar year stratification

There were 3,892 patients with 3,047 (78.3%) deaths over the study period in GOLD with a median follow up of 0.58 (IQR 0.17-1.58) years. The median survival for the whole population was 0.70 (95%CI 0.65-0.74) years with a slightly higher median survival of around 0.80 years (95%CI 0.75-0.84) in Aurum (Supplement S8). Survival after one-, five- and ten-years after diagnosis was 41.7%, 13.2% and 7.1% for both sexes. Similar observations although slightly higher were observed in Aurum (Supplement S9).

Median survival was slightly lower for females compared to males across both databases. In GOLD, females had a median survival of 0.62 (95% CI 0.56-0.69) years whereas males had median survival of 0.74 years (95% CI 0.68-0.81). Stratifying by age group, the general trend indicates median survival decreased from 40 years onwards for both databases (Supplement S10). Short term survival decreased with age from 30-39 years of age with the lowest survival observed in those aged 90 years and older (Table 2).

**Table 2:** Survival rates of PLC from 2000 to 2019 for CPRD Aurum and 2000-2021 for GOLD stratified by database and age group.

| **Age Group** | **One year survival (%)** |  | **Five-year survival (%)** |  | **Ten-year survival (%)** |  |
| --- | --- | --- | --- | --- | --- | --- |
|  | Aurum | GOLD | Aurum | GOLD | Aurum | GOLD |
| 18-29 | 62.9 (47.6 - 83.0) | 31.8 (14.2 - 70.8) | 46 (30.3-69.9) | - | - | - |
| 30-39 | 65.7 (55.7 - 77.3) | 62.5 (45.7 - 85.5) | 51.9 (41 - 65.7) | 44.2 (26.7 - 73.3) | 46.7 (34.1-63.9) | - |
| 40-49 | 60.3 (54.9 - 66.3) | 54.9 (47.1 - 64.1) | 35.8 (30 - 42.7) | 29.1 (21.5 - 39.3) | 25.5 (19.5 - 33.3) | 26.5 (19.1 - 36.8) |
| 50-59 | 53.3 (50.3 - 56.5) | 48.4 (44.3 - 52.9) | 22.4 (19.6-25.7) | 20.3 (16.8 - 24.7) | 17.1 (14.1 - 20.7) | 12.5 (9.0 - 17.4) |
| 60-69 | 50.7 (48.4 - 53.1) | 47.8 (44.9 - 51.0) | 19.1 (17.0-21.4) | 16.0 (13.6 - 18.9) | 10.3 (8.3 - 12.8) | 9.2 (6.9 - 12.4) |
| 70-79 | 42.4 (40.3 - 44.5) | 37.7 (35.1 - 40.5) | 12.4 (10.8-14.2) | 10.1 (8.3 - 12.3) | 5.3 (3.9 - 7.2) | 3.6 (2.3 - 5.7) |
| 80-89 | 33.9 (31.2 - 36.8) | 28.1 (25.0 - 31.6) | 6.9 (5.21-9.11) | 5.0 (3.3 - 7.6) | 1.9 (0.7 - 5.2) | 0.7 (0.1 - 4.3) |
| 90+ | 21.6 (15.6 - 30) | 30.3 (20.6 - 44.6) | 1.6 (0.26 - 10.2) | 3.1 (0.5 - 19.6) | - | - |

To investigate if survival has changed over time in GOLD, we stratified by calendar time of cancer diagnosis in five-year age windows. Figure 5 shows the Kaplan-Meier survival curve for the whole population, stratified by sex and calendar year for GOLD.

**Figure 5:**
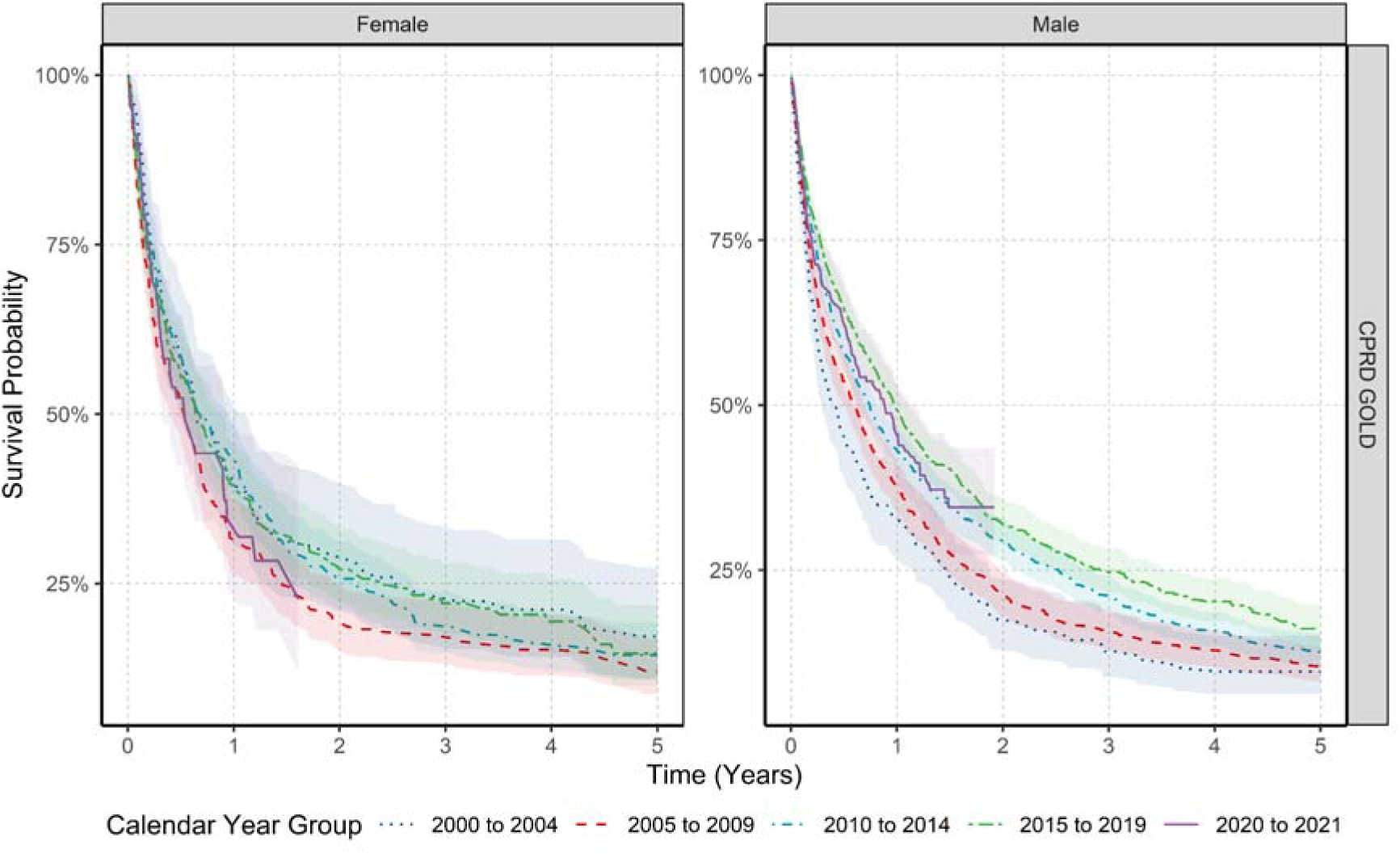
Kaplan-Meier survival curve of PLC stratified by sex and calendar year of diagnosis.

Overall, median survival increased from 0.45 years (95% CI 0.37 - 0.65) for those diagnosed in 2000-2004 to 0.912 (95% CI 0.783 - 0.997) years for those diagnosed in 2015-2019. However, when stratifying by sex this trend was only observed in males with median survival increasing from 0.427 (95% CI 0.287 - 0.597) to 0.975 (95% CI 0.862 - 1.098). There was no difference in median survival when comparing 2015-2019 with 2020-2021. Stratification by age group did not show any clear pattern in improvements in median survival over time apart from those aged 60-69 years where there was an increase in median survival comparing those diagnosed between 2005-2009 with 2015-2019 (0.63 years to 1.33 years) (Supplement S11)

In general, survival at one-year was higher in those diagnosed between 2015-2019 compared with those diagnosed between 2000-2004 with survival increasing from 35.6% (95% CI 30.5 - 41.5) to 46.7% (95% CI 43.8 - 49.9). Stratification by sex showed one-year survival increased in males only, from 33.2% (95% CI 27.2 - 40.4) to 49.3% (95% CI 45.9 - 53.1). For long term survival, there were no increases in five-year survival for the whole population and for each sex (Supplement S12). One-year survival also improved only in those aged 60-69 years of age (35.52% (95% CI 26.6 - 47.5) to 58.2% (95% CI 52.7 - 64.3)). There was no clear pattern in improvements in long term survival over calendar year for other age groups.

## DISCUSSION

This study provides a comprehensive descriptive analysis of the trends in PLC epidemiology in the UK. The incidence and prevalence of primary liver cancer (PLC) in the UK has increased from 2000 to 2021 across both sexes and age groups. Regarding survival, males had slightly higher median survival compared to females, but one-, five- and ten-year survival were similar between sexes. Short-term survival has improved only in males over the study period, no increases in long-term survival were observed overall or by sex and age strata.

Incidence rates for PLC reported here are lower but broadly in line with the National Cancer Statistics (NCS) (9.4 per 100,000 person-years 2016-2018) (6), and other global studies across high socio-demographic index countries such as the US, Australia, the Netherlands, and the UK (14,15). Slight differences are likely due to differences in study periods (2000-2021 for our data vs. 2016-2018 for NCS), possible incompleteness/misclassification in primary care records and inclusion of other subtypes of PLC such as cholangiocarcinoma in national statistics which were we excluded these subtypes in this study.

In terms of secular trends, incidence increased for both sexes across the study period with similar trends seen in other recent works from the UK, where Burton *et al* showed that PLC incidence increased from 4.4 per 100,000 in 1997 to 9.6 in 2017 in the UK, reaching a plateau since 2014 (16), and Liao *et al* revealed that age-standardised incidence rates increased over time from 2008 to 2018 (17). The present work follows up on from several Commissions which showed that standardised mortality rates from liver disease had increased 400% since 1970 and that liver disease was the third biggest cause of premature mortality in UK, while mortality by other organ diseases kept decreasing. They presented ten recommendations to reduce the burden of liver disease in the UK (18). However, the final report of the Lancet Commission in 2021 stressed the continuing increase in burden of liver disease, especially from excess alcohol consumption and obesity, which is concerning as 49% of primary liver cancers are preventable (19).

Some of the possible reasons that explain the increase in incidence of PLC in UK could be the following. Firstly, as most PLCs typically develop in patients with chronic liver disease, the changes in the comorbidity profiles of patients over recent decades are likely to play an important role in increasing PLC rates (18,20). In the last 10 years, the introduction of successful new treatments for HCV infection has generated a reduction in the prevalence of HCV-related HCCs (21). Despite this, there is an alarming rise in the prevalence of non-viral risk factors, such as high alcohol consumption and alcohol-associated liver disease (ALD) (22), which could explain the increase in PLC (23). Furthermore, data from the Health Survey for England revealed that over 60% of adults in the UK were overweight or obese in 2021, with a higher proportion of men than women (24). The increase in obesity and diabetes are also directly related to the increase of MASLD (25), which could contribute to the increase in PLC cases. The main risk factors and the underlying liver diseases (ALD, MASLD) related to PLC identified in this study are consistent with the recent data (16) showing the greater burden of non-viral risk factors in front of the classical viral etiological factors (i.e. HCV). Similar results have been described recently and the perspective is that this trend will continue over the next years (2).

Several studies have reported higher incidence rates in males compared to females in line with our results, in the UK (16,17,26) and worldwide (14,27). Higher incidence rates with increasing age are also supported by this work (28). A decline in IR in the year 2020 was observed coinciding with the COVID-19 pandemic, which bears resemblance to findings from other studies that showed a significant reduction in HCC diagnosis during the first year of the pandemic attributed to the disruption of routine healthcare (29). The pandemic also influenced the access to HCV treatment in the UK, with studies reporting a decrease of 40.2% between 2019 and 2021 (30), which could reverse some of the progress made in HCV control and may influence future PLC rates (14).

One-, five- and ten-year survival in our study are in line with UK cancer registry and Burton et al (6,16) as well as other international studies (4). Many types of cancer such as breast, prostate, stomach and colon have seen improvements in survival over recent decades in high-income countries (1,31,32), but the same tendency has not been seen in PLC, which remains one of the lowest survival rates of any cancer in the UK. As with many other cancers, liver cancer survival is significantly better when the disease is diagnosed at an early stage, however, in UK only 3 in 10 PLC are diagnosed at early stages (33) which could explain the low survival rates in this study. In this regard, since 2022, NHS England are promoting an early detection liver cancer pilot programme to help transform outcomes for PLC patients by checking for advanced fibrosis in high-risk communities (34).

Males have higher levels of disease burden for PLC compared to females. The same sex disparity trend has been observed globally (14). It has been suggested that this tendency might be driven by differences in the prevalence of HCC risk factors, a lack of adherence to follow-up/screening programs as well as epigenetics and biological factors (14,35). Despite well-established sex differences in PLC incidence, we observed slightly lower median survival and one-year survival for females in our study. Cancer Research UK statistics for the period 2015-2019, are in line with our results showing a one-year survival of 39.3% females vs 49% males in Scotland, 35.9% vs 41.8% in England, 32.5% vs 37.4% in Wales and 39.8% vs 41% in Northern Ireland, respectively (36). However, studies in Asia and the U.S. demonstrated that women with HCC present better overall survival than men likely due to better adherence to HCC surveillance (37,38). Furthermore, a study in the U.S. reported that younger women had significantly better survival than younger men; however, this survival difference was not observed in older men and women, hypothesising that sex hormones may have a role to explain sex differences in survival (39). It has also been suggested that the patterns of alcohol consumption have been underreported particularly among women, in whom the alcohol burden has increased in recent years (40), and that females are more susceptible to alcohol-induced liver injury leading to cirrhosis and an increasing risk of HCC, although the exact mechanisms are still unclear (41,42). Females are less likely to use prevention services than males, possibly related to more perceived stigma, conflicting child/family, personal needs, and financial barriers leading to delayed diagnosis (43).

Increasing age leads to a progressive physiological and metabolic reprogramming to adapt to gradual deterioration of organs and functions that can play an important role in liver carcinogenesis in the elderly (44). A UK study showed that patients with MASLD-associated HCC were older than those with other aetiologies (71.3 years vs 67.1 years) and their cancers less often detected by surveillance (45). However, elderly patients have similar success to treatments than younger patients and should be considered for all treatments after assessment of their baseline clinical status and cancer burden (46).

Encouragingly, median survival in our study has doubled from 2000-2004 to 2015-2019, reaching almost eleven months in males. Ding *et al* also showed a significant improvement in survival for HCC patients over the past three decades in the U.S., which has been attributed to advances in early diagnosis and therapeutic approaches, such as effective systemic therapy (47).

We did not observe significant improvements of survival in our study period, which could be due to numerous reasons. Firstly, despite the impressive reduction in the rates of HCV infections, the UKHSA 2023 report suggests that nearly three-quarters of people still living with chronic HCV are unaware of their infection (30). As HCV is a risk factor for PLC, this could explain no improvements in survival due to HCV leading to PLC diagnosis delays. The slight improvement in survival observed in males could be due to HCC screening programs which are aimed mainly at populations with chronic liver disease (cirrhosis), where most patients are male, which favours earlier diagnosis of PLC with more curative treatment options (4). It has been reported that liver cancers diagnosed in asymptomatic subjects within these screening programs are associated with a slight increase in survival (48). The late diagnosis of this disease, especially in women, could be related to socio-demographic factors such as scarce primary care consultation due to family and social limitations generating a delay in the diagnosis of PLC and limiting the treatment options with the consequence of lower survival. Finally, the impact of the primary prevention measures, such as hepatitis B vaccination, reducing alcohol and tobacco use, combating obesity and better control of cardio-metabolic factors and the early diagnosis using more sensitive radiologic methods, above all, could be more clearly seen in better long-term survival in the next 10 years.

The main strength of this study is the use of two large representative data sources covering the whole of the UK. CPRD GOLD covers primary care practices from England, Wales, Scotland, and Northern Ireland whereas CPRD Aurum covers England. The similarity between the results in both databases provides increased generalizability across the UK and demonstrates the robustness of our findings. Another strength of our study is the inclusion of a complete study population database for the assessment of incidence and prevalence. In contrast, cancer registry studies extrapolate the registry data to the whole population using national population statistics, potentially introducing inaccurate denominators (49). The high validity and completeness of mortality data with over 98% accuracy compared to national mortality records (50) allowed us to examine the impact of calendar time on overall survival - one of the key outcomes in cancer care.

Our study has some limitations. Firstly, primary care data without linkage to a cancer registry was used, potentially leading to misclassification and delayed recording of cancer diagnoses. However, previous validation studies have shown high accuracy and completeness of cancer diagnoses in primary care records (51). Secondly, our use of primary care records also precluded us from studying tumour histology, genetic mutations, staging of tumour at diagnosis, or cancer therapies, which can all impact PLC survival. Finally, the main risk factors for PLC, including the percentage of underlying chronic liver disease, may be underrepresented in this study, as in large population primary care databases they may be infrequently recorded, or they might be diagnosed at the same time of the PLC and therefore not included. The patterns of alcohol consumption can be also underreported or underrecognized, particularly among women, being treated for medical conditions related to alcohol use (liver disease) or unrelated to alcohol use (MASLD) (40).

In summary, the present study shows the number of people diagnosed with PLC in the UK has substantially increased in the last 20 years and that overall survival remains low. Understanding the shift in the risk factors for PLC from virus-related to non-viral liver disease requires more research and resources to manage the care of patients with ALD and MASLD among others. Although important therapeutic advances have been made and survival has slightly improved over time, over half of the patients with PLC are not alive after one year. Therefore, further progress in prevention, early detection, and public health interventions such as screening programmes and education campaigns are needed.

## Data Availability

This study is based in part on data from the Clinical Practice Research Datalink (CPRD) obtained under the University of Oxford multi-study licence from the UK Medicines and Healthcare products Regulatory Agency. The data is provided by patients and collected by the NHS as part of their care and support. The interpretation and conclusions contained in this study are those of the author/s alone. Patient level data used in this study was obtained through an approved application to the CPRD (application number 22_001843) and is only available following an approval process to safeguard the confidentiality of patient data. Details on how to apply for data access can be found at https://cprd.com/data-access.

## ABBREVIATIONS

ALD: Alcohol-associated liver disease
CPRD: Clinical Practice Research Datalink
HCC: Hepatocellular carcinoma
HCV: Hepatitis C virus
HBV: Hepatitis B virus
IR: Incidence rate
IQR: Interquartile range
MASLD: Metabolic dysfunction-associated steatotic liver disease
NAFLD: Non-alcoholic fatty liver disease
OMOP: Observational Medical Outcomes Partnership
PLC: Primary Liver Cancer
PP: Period prevalence
SNOMED CT: Systematized Nomenclature of Medicine - Clinical Terms

## ACKNOWLEDGEMENTS

None.

## Notes

**CONFLICTS OF INTEREST** Professor Daniel Prieto-Alhambra research group has received research grants from the European Medicines Agency, from the Innovative Medicines Initiative, from Amgen, Chiesi, and from UCB Biopharma; and consultancy or speaker fees (paid to his department) from Astellas, Amgen, Astra Zeneca, and UCB Biopharma. All other authors declare no conflicts of interest.

### Competing Interest Statement

Professor Daniel Prieto-Alhambra research group has received research grants from the European Medicines Agency, from the Innovative Medicines Initiative, from Amgen, Chiesi, and from UCB Biopharma; and consultancy or speaker fees (paid to his department) from Astellas, Amgen, Astra Zeneca, and UCB Biopharma. All other authors declare no conflicts of interest.

### Funding Statement

This activity under the European Health Data & Evidence Network (EHDEN) and OPTIMA has received funding from the Innovative Medicines Initiative 2 (IMI2) Joint Undertaking under grant agreement No 806968 and No. 101034347 respectively. IMI2 receives support from the European Union's Horizon 2020 research and innovation programme and European Federation of Pharmaceutical Industries and Associations (EFPIA). The sponsors of the study did not have any involvement in the writing of the manuscript or the decision to submit it for publication. Additionally, there was partial support from the Oxford NIHR Biomedical Research Centre. The corresponding author had full access to all the data in the study and had final responsibility for the decision to submit for publication. EAT is a recipient of a Joan Rodes award from the ISCII (JR20/00047) and the PI21/01995 grant from the ISCIII-Fondos Feder.

### Author Declarations

The protocol for this research was approved by the Research Data Governance (RDG) Board of the Medicine and Healthcare products Regulatory Agency database research (protocol number 22_001843).

## REFERENCES

1. Sung H, Ferlay J, Siegel RL, Laversanne M, Soerjomataram I, Jemal A, et al. Global Cancer Statistics 2020: GLOBOCAN Estimates of Incidence and Mortality Worldwide for 36 Cancers in 185 Countries. CA Cancer J Clin. 2021;71(3):209–49.

2. Llovet JM, Kelley RK, Villanueva A, Singal AG, Pikarsky E, Roayaie S, et al. Hepatocellular carcinoma. Nat Rev Dis Primers. 2021;21;7(1):6.

3. Petrick JL, Braunlin M, Laversanne M, Valery PC, Bray F, McGlynn KA. International trends in liver cancer incidence, overall and by histologic subtype, 1978-2007. Int J Cancer. 2016;1;139(7):1534–45.

4. Lepage C, Capocaccia R, Hackl M, Lemmens V, Molina E, Pierannunzio D, et al. Survival in patients with primary liver cancer, gallbladder and extrahepatic biliary tract cancer and pancreatic cancer in Europe 1999–2007: Results of EUROCARE-5. Eur J Cancer. 2015;51(15):2169–78.

5. Smittenaar CR, Petersen KA, Stewart K, Moitt N. Cancer incidence and mortality projections in the UK until 2035. Br J Cancer. 2016;25;115(9):1147-1155.

6. Cancer Research UK. Liver Cancer Statistics. Accessed 14 March 2024. https://www.cancerresearchuk.org/health-professional/cancer-statistics/statistics-by-cancer-type/liver-cancer.

7. Yang JD, Hainaut P, Gores GJ, Amadou A, Plymoth A, Roberts LR. A global view of hepatocellular carcinoma: trends, risk, prevention and management. Nat Rev Gastroenterol Hepatol. 2019;Oct;16(10):589–604.

8. Kim D, Li AA, Gadiparthi C, Khan MA, Cholankeril G, Glenn JS, et al. Changing Trends in Etiology-Based Annual Mortality From Chronic Liver Disease, From 2007 Through 2016. Gastroenterology. 2016;1163(e3).

9. Herrett E, Gallagher AM, Bhaskaran K, Forbes H, Mathur R, Staa T, et al. Data Resource Profile: Clinical Practice Research Datalink (CPRD). Int J Epidemiol. 2015;44(3):827–36.

10. Voss EA, Makadia R, Matcho A, Ma Q, Knoll C, Schuemie M, et al. Feasibility and utility of applications of the common data model to multiple, disparate observational health databases. J Am Med Inform Assoc. 2015;22(3):553–64.

11. Hripcsak G, Duke JD, Shah NH, Reich CG, Huser V, Schuemie MJ, et al. Observational Health Data Sciences and Informatics (OHDSI): Opportunities for Observational Researchers. Stud Health Technol Inform. 2015;216:574–8.

12. Gilbert J, Rao G, Schuemie M, Ryan P, Weaver J. CohortDiagnostics: Diagnostics for OHDSI Cohorts. R package version 3.2.5, https://github.com/OHDSI/CohortDiagnostics, https://ohdsi.github.io/CohortDiagnostics. 2023.

13. Raventós B, Català M, Du M, Guo Y, Black A, Inberg G, et al. IncidencePrevalence: An R package to calculate population-level incidence rates and prevalence using the OMOP common data model. Pharmacoepidemiol Drug Saf. 2024;33(1):e5717.

14. Rumgay H, Arnold M, Ferlay J, Lesi O, Cabasag CJ, Vignat J, et al. Global burden of primary liver cancer in 2020 and predictions to 2040. J Hepatol. 2022;77(6):1598–1606.

15. Liu Z, Jiang Y, Yuan H, Fang Q, Cai N, Suo C, et al. The trends in incidence of primary liver cancer caused by specific etiologies: Results from the Global Burden of Disease Study 2016 and implications for liver cancer prevention. J Hepatol. 2019;70(4):674–683.

16. Burton A, Tataru D, Driver RJ, Bird TG, Huws D, Wallace D, et al. Primary liver cancer in the UK: Incidence, incidence-based mortality, and survival by subtype, sex, and nation. JHEP Rep. 2021;19;3(2):100232.

17. Liao W, Coupland CAC, Innes H, Jepsen P, Matthews PC, Campbell C, et al. Disparities in care and outcomes for primary liver cancer in England during 2008-2018: a cohort study of 8.52 million primary care population using the QResearch database. EClinicalMedicine. 2023;11;59:101969.

18. Williams R, Aspinall R, Bellis M, Camps-Walsh G, Cramp M, Dhawan A, et al. Addressing liver disease in the UK: a blueprint for attaining excellence in health care and reducing premature mortality from lifestyle issues of excess consumption of alcohol, obesity, and viral hepatitis. The Lancet. 2014;384(9958):1953–97.

19. Williams R, Aithal G, Alexander GJ, Allison M, Armstrong I, Aspinall R, et al. Unacceptable failures: the final report of the Lancet Commission into liver disease in the UK. The Lancet. 2020;395(10219):226–39.

20. Singal AG, Kanwal F, Llovet JM. Global trends in hepatocellular carcinoma epidemiology: implications for screening, prevention and therapy. Nat Rev Clin Oncol. 2023;20(12):864–884.

21. Harris HE, Costella A, Croxford S. Hepatitis C in the UK, 2020: Working to Eliminate Hepatitis C as a Major Public Health Threat. 2020.

22. Huang DQ, Mathurin P, Cortez-Pinto H, Loomba R. Global epidemiology of alcohol-associated cirrhosis and HCC: trends, projections and risk factors. Nat Rev Gastroenterol Hepatol. 2023;20(1):37–49.

23. Karlsen TH, Sheron N, Zelber-Sagi S, Carrieri P, Dusheiko G, Bugianesi E, et al. The EASL–Lancet Liver Commission: protecting the next generation of Europeans against liver disease complications and premature mortality. The Lancet. 2022;399(10319):61–116.

24. NHS Digital, Health Survey for England, 2021 part 1. https://digital.nhs.uk/data-and-information/publications/statistical/health-survey-for-england/2021/overweight-and-obesity-in-adults. Accessed 11 January 2024.

25. Orci LA, Sanduzzi-Zamparelli M, Caballol B, Sapena V, Colucci N, Torres F, et al. Incidence of Hepatocellular Carcinoma in Patients With Nonalcoholic Fatty Liver Disease: A Systematic Review, Meta-analysis, and Meta-regression. Clin Gastroenterol Hepatol. 2022;20(2):283–292.e10.

26. Burton A, Balachandrakumar VK, Driver RJ, Tataru D, Paley L, Marshall A, et al. Regional variations in hepatocellular carcinoma incidence, routes to diagnosis, treatment and survival in England. Br J Cancer. 2022;126(5):804–814.

27. McGlynn KA, Petrick JL, El-Serag HB. Epidemiology of Hepatocellular Carcinoma. Hepatology. 2021;73 Suppl 1(Suppl 1):4–13.

28. Petrick JL, Florio AA, Znaor A, Ruggieri D, Laversanne M, Alvarez CS, et al. International trends in hepatocellular carcinoma incidence, 1978-2012. Int J Cancer. 2020;147(2):317–30.

29. Geh D, Watson R, Sen G, French JJ, Hammond J, Turner P, et al. COVID-19 and liver cancer: lost patients and larger tumours. BMJ Open Gastroenterol. 2022;9(1):e000794.

30. Hepatitis C in the UK 2023. Working to eliminate hepatitis C as a public health threat. Data to end of 2021. https://assets.publishing.service.gov.uk/media/63da9c888fa8f5187bafd5e5/hepatitis-c-in-the-UK-2023.pdf.

31. Arnold M, Rutherford MJ, Bardot A, Ferlay J, Andersson TML, Myklebust TÅ, et al. Progress in cancer survival, mortality, and incidence in seven high-income countries 1995–2014 (ICBP SURVMARK-2): a population-based study. Lancet Oncol. 2019;20(11):1493–505.

32. Allemani C, Weir HK, Carreira H, Harewood R, Spika D, Wang XS, et al. Global surveillance of cancer survival 1995–2009: analysis of individual data for 25lJ676lJ887 patients from 279 population-based registries in 67 countries (CONCORD-2). The Lancet. 2015;385(9972):977–1010.

33. Liver Cancer UK. Accessed 14 May 2024. https://livercanceruk.org/liver-cancer-information/statistics/#latediagnosis.

34. NHS England. https://www.england.nhs.uk/2023/10/nhs-mobile-testing-scheme-finds-thousands-of-new-cases-of-liver-damage/ Accessed 05 May 2024.

35. Liou WL, Tan TJ, Chen K, Goh GB, Chang JP, Tan CK. Gender survival differences in hepatocellular carcinoma: Is it all due to adherence to surveillance? A study of 1716 patients over three decades. JGH Open. 2023;24;7(5):377–386.

36. Cancer Research UK. Cancer Statistics Data Hub. https://crukcancerintelligence.shinyapps.io/CancerStatsDataHub/ Accessed 05 May 2024.

37. Liou W, Tan TJ, Chen K, Goh GB, Chang JP, Tan C. Gender survival differences in hepatocellular carcinoma: Is it all due to adherence to surveillance? A study of 1716 patients over three decades. JGH Open. 2023;7(5):377–86.

38. Phipps M, Livanos A, Guo A, Pomenti S, Yeh J, Dakhoul L, et al. Gender Matters: Characteristics of Hepatocellular Carcinoma in Women From a Large, Multicenter Study in the United States. American Journal of Gastroenterology. 2020;115(9):1486– 95.

39. Rich NE, Murphy CC, Yopp AC, Tiro J, Marrero JA, Singal AG. Sex disparities in presentation and prognosis of 1110 patients with hepatocellular carcinoma. Aliment Pharmacol Ther. 2020;52(4):701–9.

40. Szabo G. Women and alcoholic liver disease - warning of a silent danger. Nat Rev Gastroenterol Hepatol. 2018;15(5):253–254.

41. Yin M, Ikejima K, Wheeler MD, Bradford BU, Seabra V, Forman DT, et al. Estrogen is involved in early alcohol-induced liver injury in a rat enteral feeding model. Hepatology. 2000;31(1):117–23.

42. Bizzaro D, Becchetti C, Trapani S, Lavezzo B, Zanetto A, D’Arcangelo F, et al. Influence of sex in alcohol-related liver disease: Pre-clinical and clinical settings. United European Gastroenterol J. 2023;11(2):218–27.

43. Jophlin LL, Singal AK, Bataller R, Wong RJ, Sauer BG, Terrault NA, et al. ACG Clinical Guideline: Alcohol-Associated Liver Disease. Am J Gastroenterol. 2024;119(1):30–54.

44. Macias RIR, Monte MJ, Serrano MA, González-Santiago JM, Martín-Arribas I, Simão AL, et al. Impact of aging on primary liver cancer: epidemiology, pathogenesis and therapeutics. Aging (Albany NY). 2021;11;13(19):23416–34.

45. Dyson J, Jaques B, Chattopadyhay D, Lochan R, Graham J, Das D, et al. Hepatocellular cancer: The impact of obesity, type 2 diabetes and a multidisciplinary team. J Hepatol. 2014 Jan;60(1):110–7.

46. Hung AK, Guy J. Hepatocellular carcinoma in the elderly: Meta-analysis and systematic literature review. World J Gastroenterol. 2015;21(42):12197.

47. Ding J, Wen Z. Survival improvement and prognosis for hepatocellular carcinoma: analysis of the SEER database. BMC Cancer. 2021;21(1):1157.

48. Choi DT, Kum HC, Park S, Ohsfeldt RL, Shen Y, Parikh ND, et al. Hepatocellular Carcinoma Screening Is Associated With Increased Survival of Patients With Cirrhosis. Clinical Gastroenterology and Hepatology. 2019;17(5):976–987.e4.

49. Sarfati D, Blakely T, Pearce N. Measuring cancer survival in populations: relative survival vs cancer-specific survival. Int J Epidemiol. 2010;39(2):598–610.

50. Gallagher AM, Dedman D, Padmanabhan S, Leufkens HGM, Vries F. The accuracy of date of death recording in the Clinical Practice Research Datalink GOLD database in England compared with the Office for National Statistics death registrations. Pharmacoepidemiol Drug Saf. 2019;28(5):563–569.

51. Strongman H, Williams R, Bhaskaran K. What are the implications of using individual and combined sources of routinely collected data to identify and characterise incident site-specific cancers? a concordance and validation study using linked English electronic health records data. BMJ Open. 2020;20;10(8):e037719.

