## supplement for "Secular trends in the incidence, prevalence, and survival of primary liver cancer in the United Kingdom from 2000-2021: a population-based cohort study"

### **S1: Clinical codelists for PLC**

The clinical codelists used for each cancer are listed in the table below with the corresponding SNOMED concept ID, OMOP concept ID and concept description. Only diagnosis records alone were used to identify cancer outcome for this study. Different codelists were created for incident and prevalent definitions of PLC. We developed concept definitions using ATLAS, the OHDSI open-source platform (<https://github.com/OHDSI/atlas>). Clinical adjudicators reviewed the cohort definitions and associated concept sets.

| **Concept Id** | **Concept SNOMED Code** | **Concept Description** | **Code used for** |
| --- | --- | --- | --- |
| 4001171 | 109841003 | Liver cell carcinoma | Incidence and prevalence |
| 4246127 | 93870000 | Malignant neoplasm of liver | Incidence and prevalence |
| 201519 | 95214007 | Primary malignant neoplasm of liver | Incidence and prevalence |
| 4094864 | 187769009 | Primary carcinoma of liver | Incidence and prevalence |
| 4095432 | 187767006 | Malignant neoplasm of liver and intrahepatic bile ducts | Incidence and prevalence |
| 4001172 | 109843000 | Hepatoblastoma | Incidence and prevalence |
| 4003021 | 109844006 | Angiosarcoma of liver | Incidence and prevalence |
| 4099699 | 253018005 | Fibrolamellar hepatocellular carcinoma | Incidence and prevalence |
| 4166154 | 274902006 | Combined hepatocellular carcinoma and cholangiocarcinoma | Incidence and prevalence |
| 4252535 | 408646000 | Adenocarcinoma of liver | Incidence and prevalence |
| 36674832 | 770685009 | Undifferentiated carcinoma of liver and intrahepatic biliary tract | Incidence and prevalence |
| 37204022 | 787091002 | Adenocarcinoma of liver and intrahepatic biliary tract | Incidence and prevalence |
| 37311916 | 788982002 | Mesothelial carcinoma of liver | Incidence and prevalence |
| 37396736 | 716648006 | Embryonal sarcoma of liver | Incidence and prevalence |
| 4196266 | 315000005 | Metastasis from malignant tumor of liver | Prevalence only |
| 4200888 | 314963000 | Local recurrence of malignant tumor of liver | Prevalence only |

### **S2: Population attrition showing eligible patients for study from each database.**

| **N** | **Reason** | **N excluded** | **Database** |
| --- | --- | --- | --- |
| 39999011 | Starting population |  | Aurum |
| 39999011 | Missing year of birth | 0 |  |
| 39999011 | Missing sex | 0 |  |
| 34833388 | Cannot satisfy age criteria during the study period based on year of birth | 5165623 |  |
| 29190480 | No observation time available during study period | 5642908 |  |
| 29190480 | Doesn't satisfy age criteria during the study period | 0 |  |
| 25483313 | Prior history requirement not fulfilled during study period | 3707167 |  |
| 24340860 | No observation time available after applying age and prior history criteria | 1142453 |  |
| 24340860 | Starting analysis population |  |  |
| 24340860 | Estimating prevalence |  |  |
| 24340327 | Excluded due to prior event (do not pass outcome washout during study period) | 533 |  |
| 24340327 | Estimating incidence |  |  |
| 7281 | With a cancer diagnosis | 24333046 |  |
| 7103 | Cancer diagnosis not on same date as death | 178 |  |
| 7103 | Estimating survival |  |  |
| 17054819 | Starting population |  | GOLD |
| 17054819 | Missing year of birth | 0 |  |
| 17054819 | Missing sex | 0 |  |
| 15210165 | Cannot satisfy age criteria during the study period based on year of birth | 1844654 |  |
| 13978229 | No observation time available during study period | 1231936 |  |
| 13978229 | Doesn't satisfy age criteria during the study period | 0 |  |
| 12254874 | Prior history requirement not fulfilled during study period | 1723355 |  |
| 11388117 | No observation time available after applying age and prior history criteria | 866757 |  |
| 11388117 | Starting analysis population |  |  |
| 11388117 | Estimating prevalence |  |  |
| 11387938 | Excluded due to prior event (do not pass outcome washout during study period) | 179 |  |
| 11387938 | Estimating incidence |  |  |
| 3999 | With a cancer diagnosis | 11383939 |  |
| 3892 | Cancer diagnosis not on same date as death | 107 |  |
| 3892 | Estimating survival |  |  |

### **S3: Baseline characteristics of PLC patients at the time of diagnosis for CPRD Aurum.**

| **Database** | **CPRD Aurum** |
| --- | --- |
| **Number of patients** | 7,281 |
| **Sex: Male (N[%])** | 5,274 (72.40%) |
| **Age (Median [IQR])** | 70 (61 to 78) |
| **Age Groups N (%)** |  |
| 18-29 | 32 (0.40%) |
| 30-39 | 80 (1.10%) |
| 40-49 | 316 (4.30%) |
| 50-59 | 1,079 (14.80%) |
| 60-69 | 1,929 (26.50%) |
| 70-79 | 2,377 (32.60%) |
| 80-89 | 1,305 (17.90%) |
| 90+ | 163 (2.20%) |
| **Prior history, days** |  |
| median [IQR] | 6,240 (2,938 to 10,310) |
| **General conditions (any time prior)** |  |
| Atrial fibrillation | 581 (8.00%) |
| Cerebrovascular disease | 556 (7.60%) |
| Chronic liver disease | 1,860 (25.50%) |
| Chronic obstructive lung disease | 730 (10.00%) |
| Coronary arteriosclerosis | 118 (1.60%) |
| Crohn's disease | 38 (0.50%) |
| Dementia | 77 (1.10%) |
| Depressive disorder | 986 (13.50%) |
| Diabetes | 2,644 (36.30%) |
| Gastroesophageal reflux disease | 259 (3.60%) |
| Gastrointestinal hemorrhage | 905 (12.40%) |
| Heart disease | 1960 (26.90%) |
| Heart failure | 405 (5.60%) |
| Hepatitis C | 489 (6.70%) |
| HIV | 7 (0.10%) |
| Hyperlipidemia | 669 (9.20%) |
| Hypertensive disorder | 3,394 (46.60%) |
| Ischemic heart disease | 1,093 (15.00%) |
| Lesion of Liver | 2,219 (30.50%) |
| Osteoarthritis | 402 (5.50%) |
| Peripheral vascular disease | 2,038 (28.00%) |
| Pneumonia | 251 (3.40%) |
| Psoriasis | 303 (4.20%) |
| Pulmonary embolism | 360 (4.90%) |
| Renal impairment | 114 (1.60%) |
| Rheumatoid arthritis | 1,225 (16.80%) |
| Schizophrenia | 89 (1.20%) |
| Ulcerative colitis | 31 (0.40%) |
| Urinary tract infectious disease | 71 (1.00%) |
| Venous thrombosis | 987 (13.60%) |
| Visual system disorder | 469 (6.40%) |

### **S4: Age standardised Incidence by the European Standard Population for incidence rates for CPRD GOLD for PLC stratified by sex (red dotted line denotes introduction of Quality and Outcomes Framework (QOF) in 2004)**

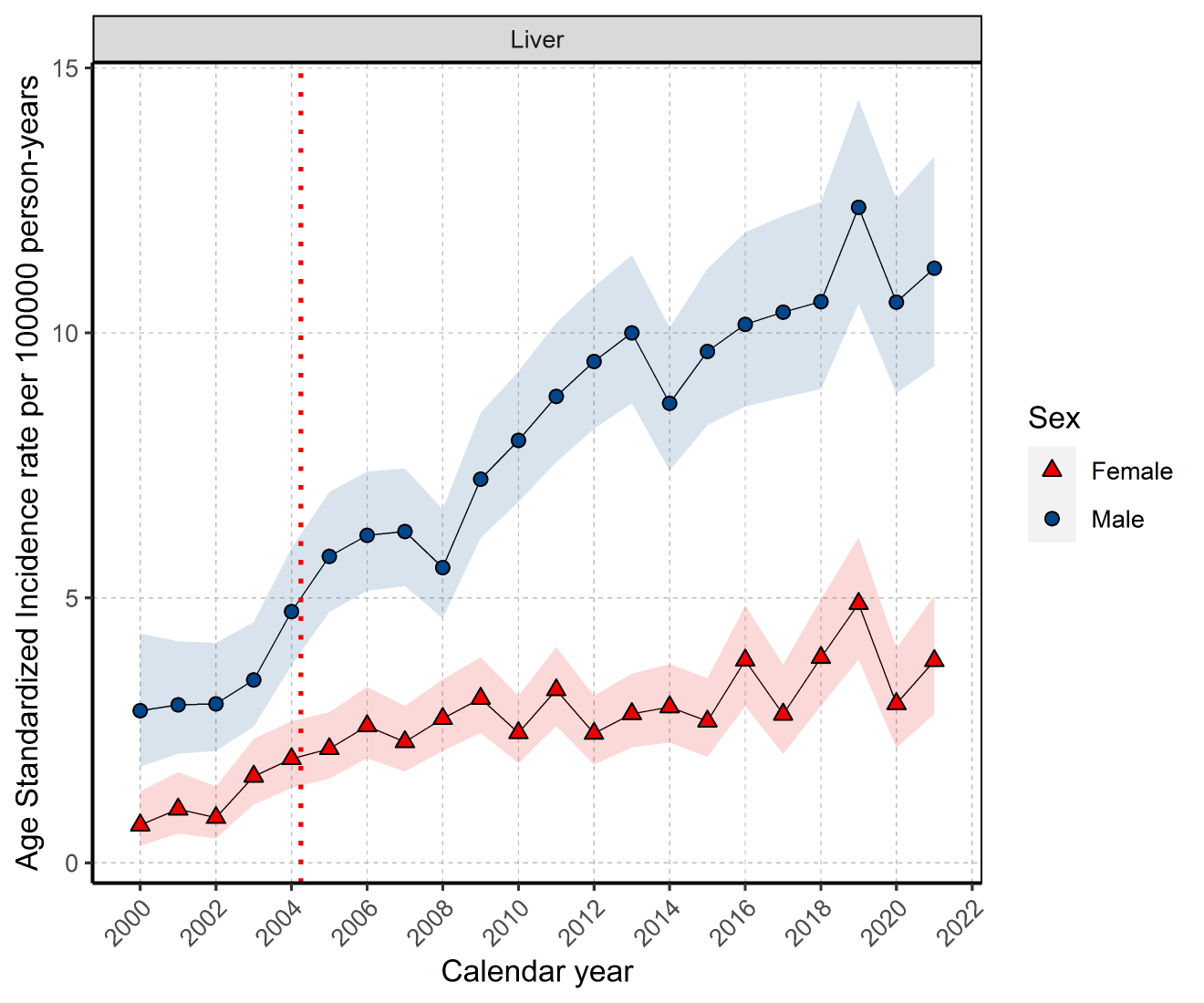

### **S5: Overall incidence rates for PLC across study period for GOLD (2000-2021) and Aurum (2000-2019) stratified by database and age group.**

| **Age Group** | **n persons** | **pys** | **n events** | **Incidence (100,000 pys)** | **Database** |
| --- | --- | --- | --- | --- | --- |
| 18 to 29 | 9,238,440 | 31,941,413 | 32 | 0.10 (0.07 to 0.14) | CPRD Aurum |
| 30 to 39 | 8,292,694 | 32,680,491 | 80 | 0.24 (0.19 to 0.30) |  |
| 40 to 49 | 6,516,214 | 32,926,479 | 316 | 0.96 (0.86 to 1.07) |  |
| 50 to 59 | 5,439,374 | 28,680,044 | 1,079 | 3.76 (3.54 to 3.99) |  |
| 60 to 69 | 4,173,598 | 22,533,850 | 1,929 | 8.56 (8.18 to 8.95) |  |
| 70 to 79 | 3,125,160 | 16,299,264 | 2,377 | 14.6 (14.0 to 15.2) |  |
| 80 to 89 | 1,954,123 | 8,861,283 | 1,305 | 14.7 (13.9 to 15.6) |  |
| 90 + | 667,527 | 2,273,344 | 163 | 7.17 (6.11 to 8.36) |  |
| 18 to 29 | 3,871,125 | 16,018,603 | 14 | 0.09 (0.05 to 0.15) | CPRD GOLD |
| 30 to 39 | 3,682,421 | 15,107,603 | 27 | 0.18 (0.12 to 0.26) |  |
| 40 to 49 | 3,247,010 | 16,117,751 | 138 | 0.86 (0.72 to 1.01) |  |
| 50 to 59 | 2,886,150 | 14,727,024 | 560 | 3.80 (3.49 to 4.13) |  |
| 60 to 69 | 2,294,954 | 11,908,187 | 1,071 | 8.99 (8.46 to 9.55) |  |
| 70 to 79 | 1,684,951 | 8,430,060 | 1,337 | 15.86 (15.02 to 16.73) |  |
| 80 to 89 | 1,026,008 | 4,434,032 | 783 | 17.66 (16.44 to 18.94) |  |
| 90 + | 323,237 | 966,781 | 69 | 7.14 (5.55 to 9.03) |  |

pys: person years

### **S6: Annualised incidence rates for PLC from 2000 to 2021 stratified by database, sex and age group.**

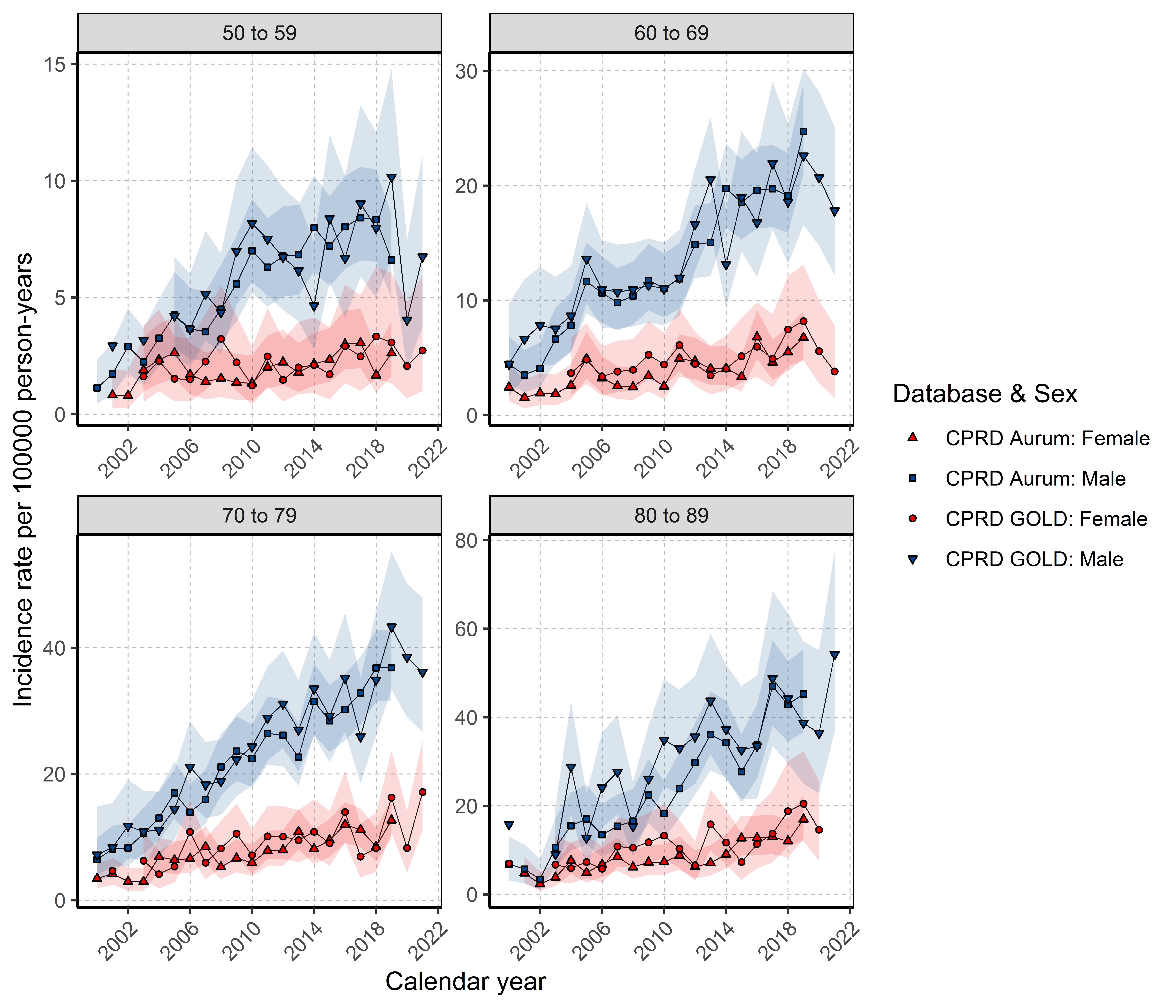

### **S7: Annualised prevalence for PLC from 2000 to 2021 stratified by database, sex and age group.**

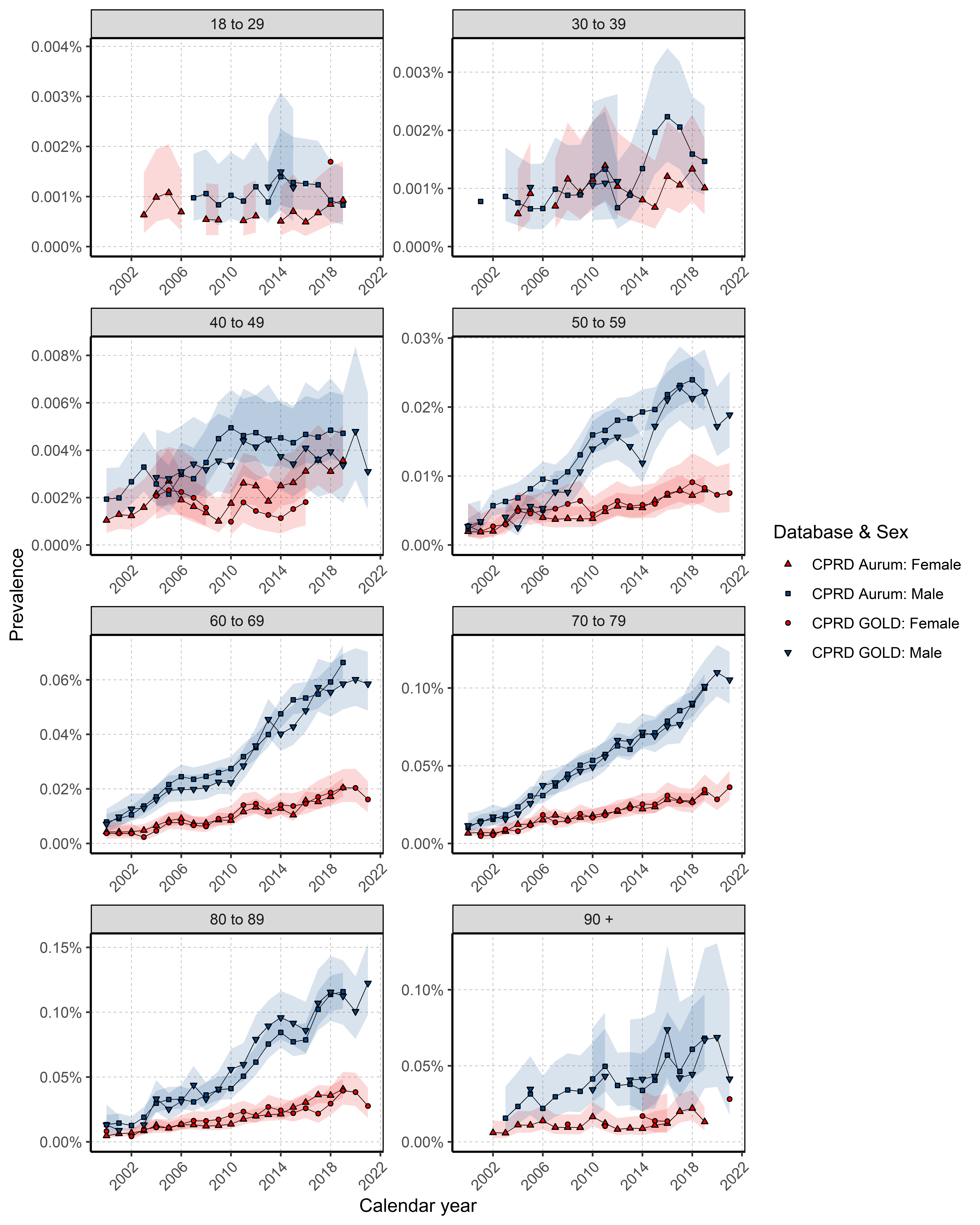

### **S8: Kaplan-Meier survival curve of PLC stratified by database and sex**

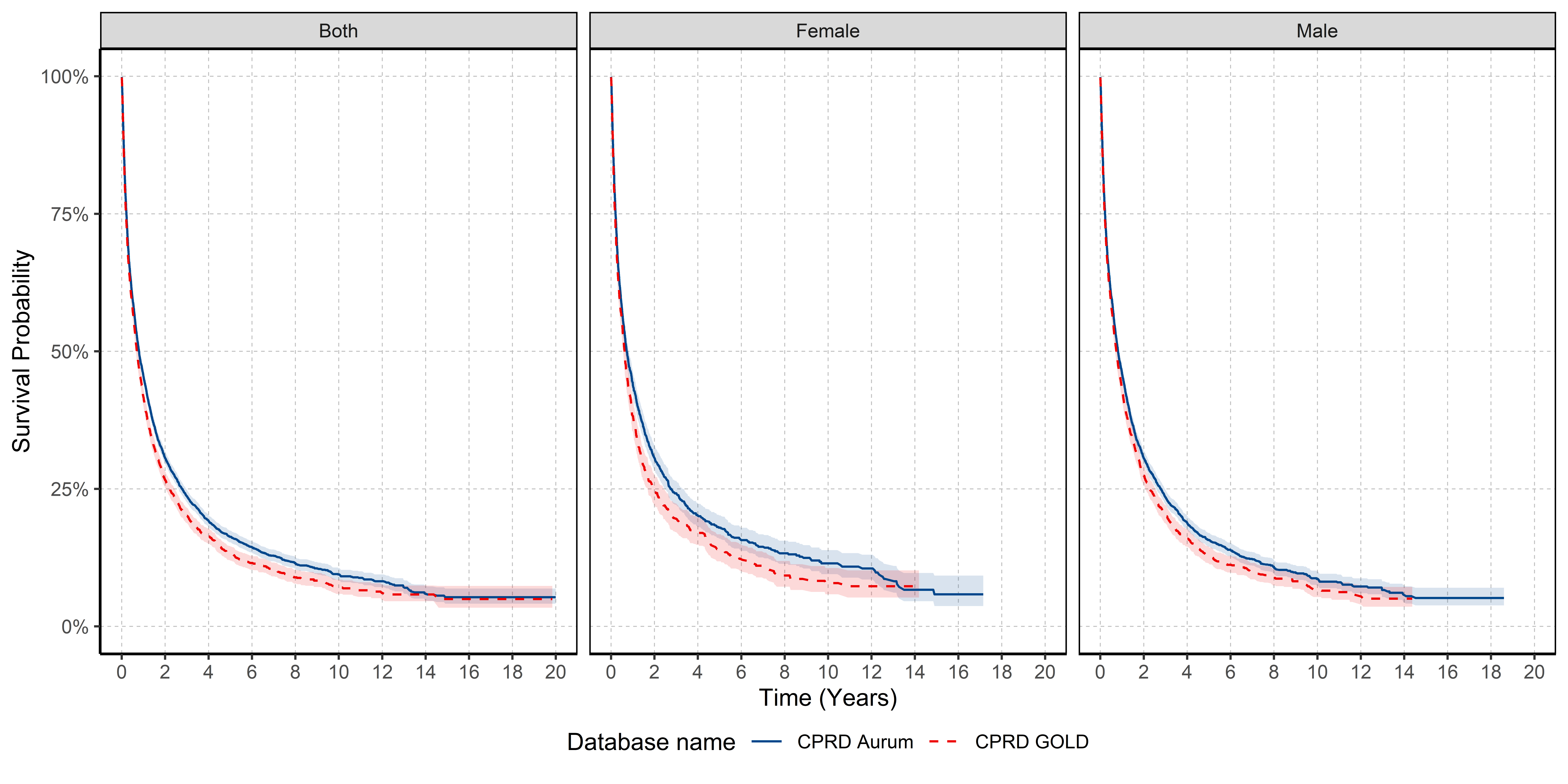

### **S9: Survival (%) after 1, 5 and 10 years after PLC diagnosis stratified by database and sex.**

| **Time** | **Sex** | **% Survival (95% CI)** | **Database** |
| --- | --- | --- | --- |
| 1 | Male | 45.9 (44.5 - 47.4) | Aurum |
| 5 |  | 15.7 (14.5 – 17.0) |  |
| 10 |  | 8.6 (7.4 - 10.0) |  |
| 1 | Female | 44.3 (42.1 - 46.7) |  |
| 5 |  | 17.9 (16.0 - 20.2) |  |
| 10 |  | 11.5 (9.5 - 13.8) |  |
| 1 | Male | 43.0 (41.1 - 44.9) | GOLD |
| 5 |  | 12.8 (11.4 - 14.5) |  |
| 10 |  | 6.7 (5.3 - 8.5) |  |
| 1 | Female | 38.4 (35.5 - 41.5) |  |
| 5 |  | 14.1 (11.8 - 16.7) |  |
| 10 |  | 7.9 (5.8 - 10.6) |  |

CI: confidence interval

### **S10: Median survival stratified by database and age group for whole population.**

| **Age Group** | **Median survival in years (95% CI)** | **n persons** | **n events** | **Database** |
| --- | --- | --- | --- | --- |
| 18 to 29 | Not achieved | 32 | 16 | Aurum |
| 30 to 39 | Not achieved | 79 | 37 |  |
| 40 to 49 | 2.059 (1.399 - 2.834) | 308 | 180 |  |
| 50 to 59 | 1.153 (1.010 - 1.333) | 1055 | 723 |  |
| 60 to 69 | 1.027 (0.925 - 1.155) | 1892 | 1352 |  |
| 70 to 79 | 0.704 (0.646 - 0.780) | 2330 | 1805 |  |
| 80 to 89 | 0.509 (0.435 - 0.597) | 1254 | 992 |  |
| 90 + | 0.244 (0.167 - 0.372) | 153 | 128 |  |
| 18 to 29 | Not achieved | 14 | 10 | GOLD |
| 30 to 39 | Not achieved | 26 | 13 |  |
| 40 to 49 | 1.383 (0.934 - 2.253) | 134 | 84 |  |
| 50 to 59 | 0.986 (0.808 - 1.175) | 544 | 393 |  |
| 60 to 69 | 0.936 (0.821 - 1.060) | 1050 | 796 |  |
| 70 to 79 | 0.632 (0.580 - 0.717) | 1305 | 1065 |  |
| 80 to 89 | 0.416 (0.350 - 0.496) | 757 | 638 |  |
| 90 + | 0.312 (0.211 - 0.758) | 62 | 48 |  |

### **S11: Median survival stratified by calendar year and sex for CPRD GOLD.**

| **Sex** | **Calendar Year** | **Median Survival in Years (95% CI)** | **Records (n)** | **Events (n)** |
| --- | --- | --- | --- | --- |
| Both | 2000 to 2004 | 0.496 (0.370 - 0.652) | 306 | 254 |
|  | 2005 to 2009 | 0.572 (0.498 - 0.660) | 914 | 771 |
|  | 2010 to 2014 | 0.739 (0.635 - 0.810) | 1241 | 987 |
|  | 2015 to 2019 | 0.912 (0.783 - 0.997) | 1079 | 779 |
|  | 2020 to 2021 | 0.843 (0.572 - 0.975) | 352 | 177 |
| Female | 2000 to 2004 | 0.652 (0.487 - 1.133) | 99 | 76 |
|  | 2005 to 2009 | 0.531 (0.402 - 0.663) | 301 | 242 |
|  | 2010 to 2014 | 0.679 (0.564 - 0.956) | 329 | 260 |
|  | 2015 to 2019 | 0.684 (0.501 - 0.914) | 299 | 214 |
|  | 2020 to 2021 | 0.539 (0.326 - 0.936) | 90 | 51 |
| Male | 2000 to 2004 | 0.427 (0.287 - 0.597) | 207 | 178 |
|  | 2005 to 2009 | 0.602 (0.485 - 0.701) | 613 | 529 |
|  | 2010 to 2014 | 0.745 (0.635 - 0.835) | 912 | 727 |
|  | 2015 to 2019 | 0.975 (0.862 - 1.098) | 780 | 565 |
|  | 2020 to 2021 | 0.890 (0.616 - 1.180) | 262 | 126 |

CI: confidence interval

### **S12: Survival (%) after 1 and 5 years after PLC diagnosis stratified by sex and calendar year for CPRD GOLD.**

| **Calendar Year** | **Time (years)** | **% Survival (95% CI)** | **Sex** |
| --- | --- | --- | --- |
| 2000 to 2004 | 1 | 35.57 (30.47 - 41.51) | Both |
| 2005 to 2009 |  | 35.34 (32.32 - 38.65) |  |
| 2010 to 2014 |  | 43.26 (40.53 - 46.17) |  |
| 2015 to 2019 |  | 46.74 (43.75 - 49.92) |  |
| 2020 to 2021 |  | 42.66 (36.85 - 49.38) |  |
| 2000 to 2004 |  | 40.57 (31.69 - 51.93) | Female |
| 2005 to 2009 |  | 31.81 (26.78 - 37.78) |  |
| 2010 to 2014 |  | 43.34 (38.19 - 49.18) |  |
| 2015 to 2019 |  | 39.65 (34.19 - 45.98) |  |
| 2020 to 2021 |  | 34.57 (24.26 - 49.25) |  |
| 2000 to 2004 |  | 33.18 (27.24 - 40.41) | Male |
| 2005 to 2009 |  | 36.99 (33.32 - 41.05) |  |
| 2010 to 2014 |  | 43.23 (40.07 - 46.64) |  |
| 2015 to 2019 |  | 49.33 (45.85 - 53.08) |  |
| 2020 to 2021 |  | 45.52 (38.82 - 53.38) |  |
| 2000 to 2004 | 5 | 12.01 (8.67 - 16.64) | Both |
| 2005 to 2009 |  | 10.83 (8.86 - 13.23) |  |
| 2010 to 2014 |  | 13.10 (11.15 - 15.40) |  |
| 2015 to 2019 |  | 15.64 (12.99 - 18.82) |  |
| 2000 to 2004 |  | 17.18 (10.78 - 27.36) | Female |
| 2005 to 2009 |  | 12.07 (8.62 - 16.90) |  |
| 2010 to 2014 |  | 14.44 (10.82 - 19.28) |  |
| 2015 to 2019 |  | 14.64 (9.86 - 21.76) |  |
| 2000 to 2004 |  | 9.56 (6.10 - 14.98) | Male |
| 2005 to 2009 |  | 10.32 (8.05 - 13.23) |  |
| 2010 to 2014 |  | 12.55 (10.33 - 15.26) |  |
| 2015 to 2019 |  | 16.07 (13.05 - 19.80) |  |
